# Nationwide Prediction of Missed and Cancelled Appointments Using Real-World EHR Data

**DOI:** 10.64898/2026.04.08.26349942

**Authors:** Seyedeh Azadeh Miran, Yan Cheng, Charles Faselis, Cynthia Brandt, Sean Vasaitis, LaQuandra Nesbitt, Linda Zanin, Senait Tekle, Ali Ahmed, Stuart J. Nelson, Qing Zeng-Treitler

**Author notes:** **Correspondence:** Dr. Qing Zeng-Treitler, Department of Clinical Research and Leadership School of Medicine and Health Sciences, George Washington University, 2600 Virginia Avenue NW, Suite 300 Washington, DC 20037, USA.

## Abstract

**Objectives:** To develop and evaluate predictive models for unused outpatient appointments (missed or cancelled) using a large national electronic health record (EHR) repository in the United States.

**Design:** Retrospective observational study using machine learning and statistical modeling.

**Setting:** A U.S. national electronic health record repository (Cerner Real World Database) covering healthcare encounters from 2010 to 2025.

**Participants:** Adult patients aged ≥18 years with routine outpatient encounters recorded in the database. One outpatient appointment with a known status was randomly selected per patient, resulting in a final analytic sample of **5,699,861 encounters**.

**Primary and Secondary Outcome Measures:** The primary outcome was whether the index outpatient appointment was attended or unused (missed or cancelled). Model performance was evaluated using area under the receiver operating characteristic curve (AUC), sensitivity, and specificity.

**Methods:** Predictors included patient characteristics (demographics and insurance type), appointment characteristics (day, time, season, and urbanicity), prior cancellation rate, and time gap between the index appointment and the previous visit. We compared the predictive performance of two machine learning models (random forest classifier and extreme gradient boosting (XGBoost)) with logistic regression. An explainable AI analysis of feature impact was performed on the final XGBoost model.

**Results:** Among **5,699,861 outpatient encounters**, **3,650,715 (64.0%)** were attended and **2,049,146 (36.0%)** were unused. XGBoost achieved the best predictive performance on the test dataset (AUC = 0.95), followed by random forest (AUC = 0.92) and logistic regression (AUC = 0.89). Feature impact score analysis revealed highly non-linear associations between predictors and the risk of unused appointments at the individual level.

**Conclusions:** Unused outpatient appointments can be accurately predicted using routinely available EHR data. Integrating predictive models into scheduling workflows may improve healthcare efficiency and optimize appointment management.

**Article Summary:** Strengths and limitations of this study

- This study used one of the largest national electronic health record datasets to develop predictive models for unused outpatient appointments.
- Multiple modeling approaches, including logistic regression and machine learning methods (random forest and XGBoost), were compared to evaluate predictive performance.
- An explainable artificial intelligence method was applied to quantify feature impact and improve model interpretability.
- The retrospective design and reliance on routinely collected EHR data may introduce data quality limitations and unmeasured confounding.
- The database did not distinguish clearly between cancelled appointments and no-shows.

## Introduction

Unused patient appointments, including both no-shows and cancellations by providers and patients, are a pervasive challenge in outpatient care. Reported no-show rates vary widely from 5% to over 30% depending on specialty, location, and patient population^1–3^. According to Tebra’s scheduling survey in 2023, 68% of the 204 surveyed providers reported that they cancelled or rescheduled appointments 1-10 times per months; and 59% of 1,075 patients reported that they had canceled or no-showed in the past 12 months^4^. While no-shows and cancellations are distinct phenomena, they both pose a challenge for healthcare providers, as they can disrupt clinic schedules, incur time and financial costs, and interfere with timely patient care or disease management^5^. In the U.S., the financial toll of unkept appointments is estimated at $150 billion annually^6^. The consequences are particularly serious in low-resource and safety-net settings, where even modest inefficiencies can compromise access for the populations being served.

Multiple studies have identified environmental and demographic factors driving no-show and cancellation rates. For example, it has been reported that Medicaid patients are at higher risk, with odds ratios ranging from 1.5 to over 4 compared with patients with commercial insurance^6^. Additional risk factors include minority status, low household income, younger age, unemployment, housing instability, and longer lead times between scheduling and appointment^6–10^. Moreover, clinical and appointment-related variables such as prior history of missed visits, evening or weekend appointments, and distance to clinic further influence attendance patterns ^4,11–13^. There is, however, less literature evidence on risk factors associated with appointment cancellation and rescheduling, even though cancellations close to appointment time can have similar effects as no-shows. Tebra’s scheduling survey identified the top reasons for patient cancelling appointments including rescheduling and no-show as work conflicts, followed by sickness, transportation issues, family emergencies, personal emergencies, weather conditions, anxiety about appointment, and social engagements^5^.

A small number of studies have developed predictive models for no-show using the risk factors, with the goal to support operational decision-making. Prior approaches include statistical models (e.g., logistic regression, LASSO) and machine learning (ML) methods such as decision trees, random forests, and gradient boosting^14,15^. Reported accuracies varied, with the area-under-curves (AUCs) ranging from 0.70 to 0.89 depending on setting^16–18^. However, many prior studies have been limited by small sample size, regional or specialty-specific cohorts, and lack of model testing in large national datasets.

Our study addresses these gaps by using the largest national dataset reported to date, containing over 80 million routine outpatient care encounters across the U.S., to develop and test predictive models of unused appointments (including no-show). We compared traditional statistical models with advanced ensemble machine learning approaches and applied a novel explainable AI approach to analyze the trained machine learning model.

## Methods

### Data Source

We used data from the Cerner Real World Database spanning 2010-2025, a comprehensive U.S. national electronic health record (EHR) repository by Oracle Health^19^.

### Study Design

The study cohort was comprised of adults aged 18 years or older identified in the Cerner database. Because patients under 18 typically do not assume full responsibility of their care, they were excluded from the analysis. Since each patient may have both used and unused appointments, the analysis was conducted at the appointment level.

To ensure independence between training, validation, and testing datasets and to minimize potential selection bias and data leakage, one outpatient encounter with a known status was randomly selected per patient. This approach also reduced the overrepresentation of frequent users. Additionally, we required that each patient have at least one prior encounter preceding the selected one to obtain historical data of appointment use. The randomly selected encounter was defined as the index appointment.

### Outcome Variable

The outcome variable was the status of the index appointment, which was categorized as “used” (including complete, patient checking out, in progress, and confirmed present) or “unused” (including rescheduled and left-before-seen). Unfortunately, the Cerner database does not clearly differentiate cancelled appointments from unkept (no-show) appointments.

### Predictor Variables

Predictors were drawn from both patient and appointment characteristics. Patient characteristics included sex, age, ethnicity and race, and insurance type. Appointment characteristics included day of week, time of day, season and urbanicity. Time of day was categorized as early morning (before 8:00 AM), morning (8:00 AM–12:00 PM), midday (12:00 PM–2:00 PM), afternoon (2:00 PM–6:00 PM), and evening (after 6:00 PM). Two additional derived variables were created for modeling unused appointments: (1) prior cancellation rate, defined as the proportion of unused appointments among all prior encounters for a given patient up to (but excluding) the index appointment; and (2) time since the previous visit, categorized as <1 week, 1–2 weeks, 2 weeks–1 month, 1–3 months, 3–6 months, 6–12 months, and >1 year. The date when appointments were scheduled was not available in the database; therefore, the interval between scheduling and the appointment could not be included as a predictor. Age and prior cancellation rate were modeled as continuous variables.

### Statistical Analysis

#### Univariate Analysis

We tested all predictors individually for relationships with the appointment use status using a Pearson’s chi-square test for categorical variables and the Mann–Whitney U test for the continuous variables, as these variables were not normally distributed.

#### Prediction Model Development

We randomly split the dataset into three subsets: 63% for training, 27% for validation, and 10% for testing. The training subsets were then used to develop the missed appointment prediction models using three algorithms:

Logistic regression: We utilized a logistic regression method to develop the prediction model. Variable selection was performed via bidirectional stepwise selection, entering predictors with p < .01 and removing predictors with p > .05. A more stringent entry threshold (p < .01) was chosen to reduce the likelihood of spurious associations given the large sample size, where even very small effects may reach conventional statistical significance at p < .05. This approach ensured that only predictors with stronger and more stable associations were retained, thereby improving the robustness of the model and minimizing overfitting. The final model included all retained predictors and was fitted on the training data. Model coefficients, standard errors, and Wald p-values were estimated.

Random Forest Classifier: We implemented a Random Forest classifier, with optimal hyperparameters determined through a randomized search with 3-fold cross-validation (RandomizedSearchCV). This search explored 30 random combinations^20^, maximizing the area under the receiver operating characteristic curve (ROC AUC). The hyperparameter search space included: number of trees (100, 250, 500), maximum depth (5,10,15), minimum samples per split (2, 5, 10), minimum samples per leaf: (1, 2, 4), and maximum number of features to consider when looking for the best split (“sqrt”, “log2”, all features for every split). A fixed random seed was applied for reproducibility. Hyperparameter tuning was performed on the training set, and the model with the best performance was selected based on its cross-validated ROC AUC score. The validation set was used to assess generalization performance prior to final testing, which was conducted on the held-out test set.

Extreme Gradient Boosting (XGBoost): We implemented an XGBoost classifier to model the outcome, with optimal hyperparameters selected through RandomizedSearchCV. The search explored 30 random combinations, maximizing the ROC AUC. This approach balances computational efficiency with effective coverage of the hyperparameter space, as supported by prior work^20^. The hyperparameter search space included: learning rate (0.01, 0.03, 0.05, 0.1), number of trees (100, 250, 500, 1000), maximum depth (3, 5, 7, 9), regularization parameters (alpha: 0, 0.1, 1, 5; lambda: 1, 5, 10), and minimum child weight (1, 3, 5). Model training incorporated early stopping based on log loss evaluated on a validation set, halting if performance did not improve for 10 consecutive rounds ^21^. The model with the best performance was selected based on cross-validated ROC AUC and retrained on the combined training and validation sets using the optimal configuration. Early stopping was disabled during final training to allow full fitting. Final evaluation was conducted on the held-out test set.

#### Model Evaluation

The testing set was used for performance evaluation and assessing generalizability. AUC, sensitivity, and specificity were used to compare model performance.

### Feature Impact Score Analysis

Feature (predictor) impact score analysis was performed on the final XGBoost model to quantify the relative contribution of each predictor to appointment cancellation. This feature impact score method by Shao^22^ is a model-agnostic approach that quantified each feature’s contribution by comparing individual-level model predictions to reference baseline values. For continuous variables, the median was used as the reference value, and for binary (encoded categorical) variables, 0 was used^22^. The standard deviation (SD) of the impact scores indicates the degree of heterogeneity in feature impact, with larger values suggesting greater heterogeneity or non-linear associations. Positive impact scores indicate that higher values of the feature were associated with a greater likelihood of unused appointment, whereas negative scores suggest a protective effect against unuse. This impact score analysis allows us to identify impactful predictors and characterize variability in their effects across patients.

### Missing data handling

Missing or null values for insurance type were categorized as “unknown” to preserve sample size, as reported in Table 1. No other variables had meaningful missing data.

**Table 1.** Distribution of Patient, Visits and Clinical/behavioral Characteristics with Appointment Adherence.

| Characteristics | Attended<br>n = 3,650,715 | Cancelled<br>n = 2,049,146 |
| --- | --- | --- |
|  | % or Mean (SD) | % or Mean (SD) |
| **Sex** |  |  |
| Female | 56.5% | 58.0% |
| Male | 43.5% | 42.0% |
| **Race** |  |  |
| White | 71.7% | 61.7% |
| Unknown | 10.7% | 10.4% |
| Other | 8.0% | 14.6% |
| Black | 7.9% | 7.6% |
| Asian | 1.7% | 5.7% |
| **Ethnicity** |  |  |
| Non-Hispanic | 54.2% | 47.1% |
| Hispanic or Latino | 17.6% | 33.3% |
| Unknown | 28.2% | 19.6% |
| **Insurance Type** |  |  |
| Commercial | 16.1% | 19.5% |
| Self-pay | 6.4% | 6.9% |
| Medicare | 10.0% | 6.3% |
| Medicaid | 9.1% | 2.6% |
| Other | 3.2% | 1.1% |
| Unknown | 55.2% | 63.6% |
| **Appointment Time** |  |  |
| Early Morning (before 8AM) | 22.7% | 12.2% |
| Morning (8-12 AM) | 7.1% | 5.3% |
| Midday (12-2 PM) | 17.0% | 9.3% |
| Afternoon (2-6 PM) | 30.8% | 34.9% |
| Evening (after 6 PM) | 22.4% | 38.3% |
| **Appointment Weekday** |  |  |
| Monday | 19.0% | 19.7% |
| Tuesday | 20.1% | 21.3% |
| Wednesday | 19.5% | 19.8% |
| Thursday | 19.0% | 17.6% |
| Friday | 17.0% | 15.3% |
| Saturday | 3.4% | 3.3% |
| Sunday | 2.0% | 3.0% |
| **Appointment Season** |  |  |
| Spring | 25.9% | 25.2% |
| Summer | 26.8% | 24.7% |
| Fall | 23.6% | 24.6% |
| Winter | 23.7% | 25.5% |
| **Time Gap Between the Last Visit Before the Index Appointment and the Index Appointment** |  |  |
| Within a week | 64.3% | 31.2% |
| Between 1 and 2 weeks | 5.4% | 10.2% |
| Between 2 weeks and 1 month | 6.7% | 14.2% |
| Between 1 and 3 months | 8.1% | 18.6% |
| Between 3 and 6 months | 4.5% | 9.4% |
| Between 6 months and a year | 4.3% | 7.1% |
| More than a year | 6.7% | 9.3% |
| **Urbanicity** |  |  |
| Rural | 15.7% | 3.4% |
| Urban | 81.9% | 95.0% |
| Unknown | 2.4% | 1.6% |
| **Age (years)** | 45.4 (23.8) | 39.0 (24.3) |
| **Prior cancellation rate (measured as a percentage)** | 0.2% (3.1%) | 9.7% (16.7%) |

Note: All comparisons were statistically significant with $p < 0.001$ . Pearson's chi-square test was used for categorical variables, and the Mann–Whitney U test was used for continuous variables.

### Software

The Python (Version 3.4.3) programming language was utilized for all data processing, statistical computing and tests.

We used the STROBE reporting guideline^23^ to guide the reporting of this observational study and completed the STROBE checklist^24^ during manuscript preparation; the checklist is provided as supplementary file 1.

### Ethics approval and informed consent

This study used de-identified data from the Cerner Real World Database (Oracle Health). As the data were de-identified and collected for routine clinical care, informed consent was not required. The study was deemed exempt from institutional review board (IRB) review.

## Results

### Cohort and Encounter Selection

Between January 2010 and January 2025, there were 1,683,393,353 encounters with 86,518,596 unique patients (**Figure 1**). A total of 319,320,022 encounters were excluded due to inactive or invalid status (e.g., none, inactive, transitory). A total of 1,271,630,395 encounters were further excluded due to the lack of a clear use status (e.g., discharge-action, active, closed, suspended), since our study focused on those explicitly labeled as “used” (including complete, patient checking out, in progress, and confirmed present) or “unused” (including rescheduled and left-before-seen). Encounters cancelled by health services (<1%) were also excluded because the rate was too low to be reliable. In total, there were 80,111,535 used encounters and 12,331,401 unused encounters remaining.

**Figure 1:**
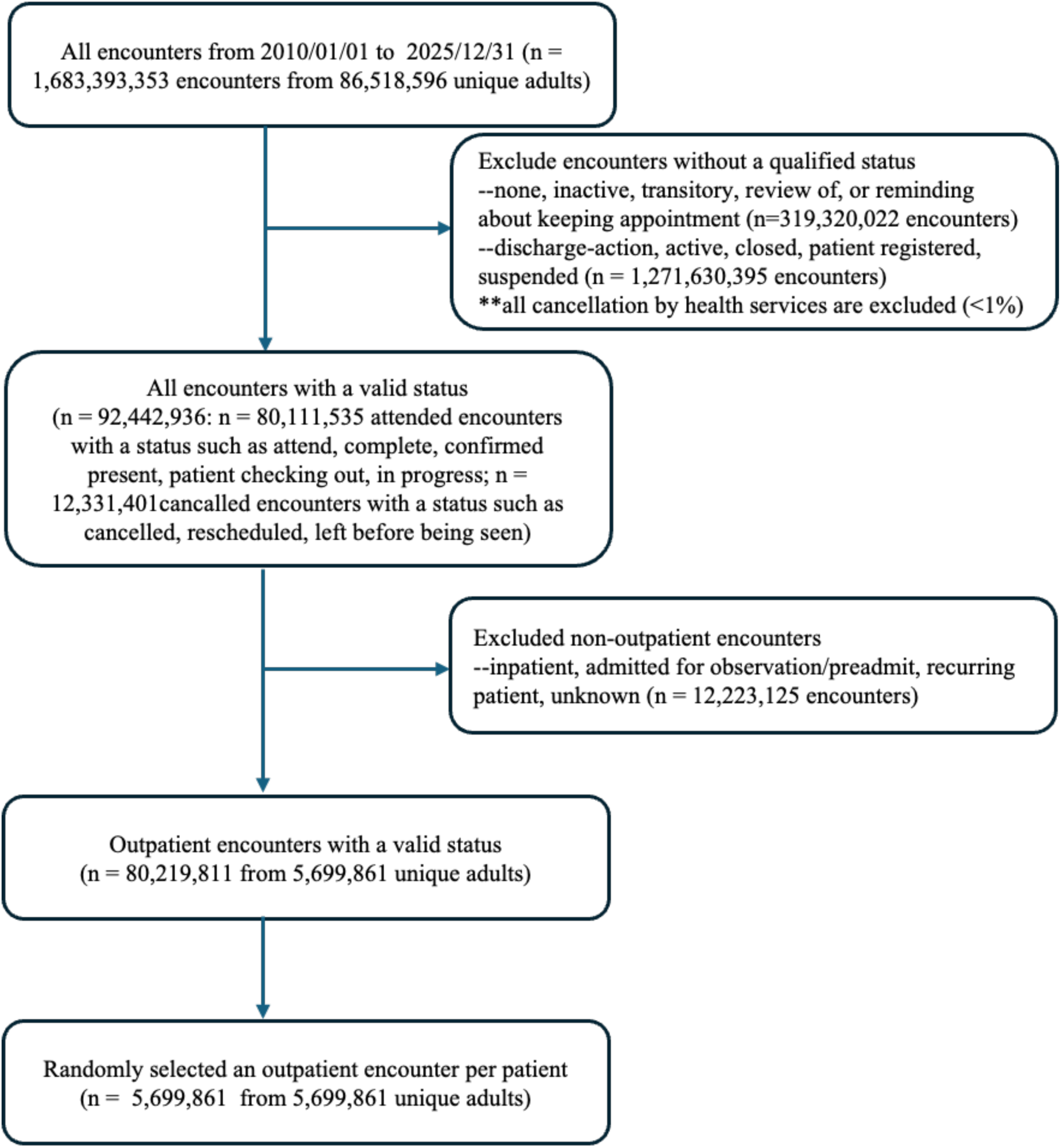
Flowchart of Cohort Selection Process.

We further restricted the data to routine outpatient visits, excluding encounters classified as hospital-based or preplanned (e.g., inpatient, admitted for observation, preadmit, or unknown visit types; approximately 12,223,125 encounters). After applying these criteria, the final cohort included 80,219,811 encounters from 5,699,861 unique patients with routine outpatient care visits, from which we selected one outpatient per patient for the predictive modeling.

### Characteristics and Appointment Cancellation

The final dataset included 5,699,861 unique patients with 3,650,715 (64.0%) used and 2,049,146 (36.0%) unused appointments (**Table 1**). Comparative analyses of patient characteristics from the **Table 1** shows that a slightly higher proportion of females did not use the appointment (58.0% vs. 56.5%, p<0.001). White patients were more likely to keep appointments (71.7% vs. 61.7%, p<0.001). Asian and “Other” race patients had high rates of unused appointments; Hispanic or Latino patients had a much higher rate of unused appointments (33.3% vs. 17.6%, p<0.001) than non-Hispanic.

Commercial insurance was more common among the unused appointments (19.5% vs. 16.1%, p<0.001). Medicaid patients had much lower proportions of unused (2.6% vs. 9.1%, p<0.001). “Unknown insurance” was disproportionately higher in the unused group (63.6% vs. 55.2%, p<0.001).

Evening appointments had the highest proportions of unused appointments (38.3% vs. 22.4%, p<0.001). Early morning and midday appointments were more likely to be attended. Weekday distribution differences are small, but Sunday was slightly more likely to be unused (3.0% vs. 2.0%, p<0.001). Seasonal differences were statistically significant but not very large. Winter appointments had a somewhat higher proportion of unused (25.5% vs. 23.7%, p<0.001). Patients seen within a week before the index appointment were much more likely to attend (64.3% vs. 31.2%, p<0.001). Those with longer gaps were more likely to not be used.

In addition, a much higher proportion of rural patients attended appointments (15.7% vs. 3.4%, p<0.001). Patients who did not use the appointment tended to be younger (mean 39.0 vs. 45.4 years, p<0.001). The prior rate of not using an appointment was dramatically higher in those who did not use the index appointment (9.7% vs. 0.2%, p<0.001).

### Model Fitting

**Figure 2** summarizes the performance of three predictive modeling approaches (logistic regression, random forest, and XGBoost) on test datasets. All models demonstrated strong discriminatory power, with logistic regression achieving an AUC of ∼0.89, random forest ∼0.92, and XGBoost ∼0.95. It indicates that all three models performed well above chance, with XGBoost clearly providing the best discrimination between used and unused appointments.

**Figure 2:**
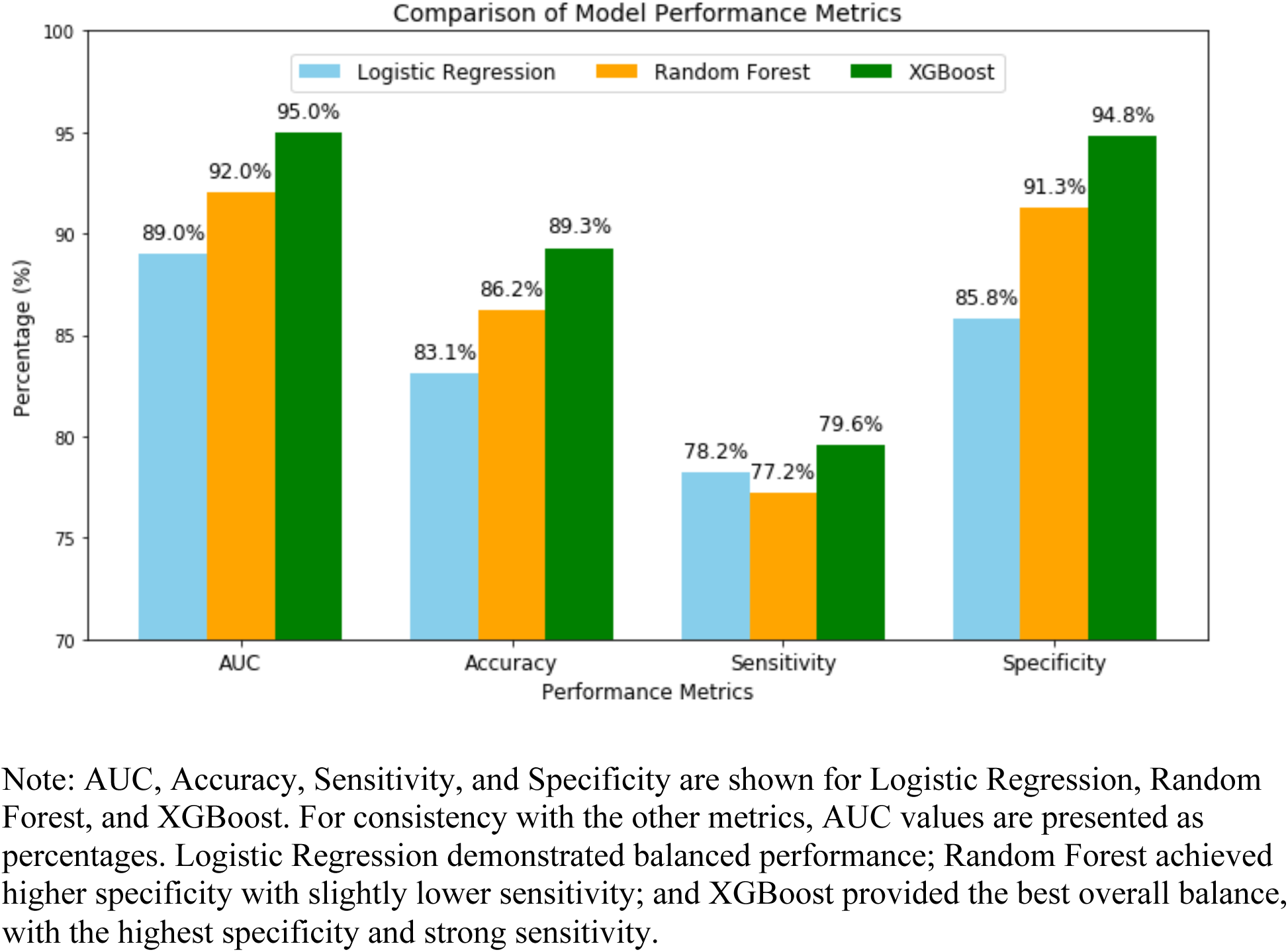
Comparative model performance metrics.

Model accuracy followed a similar trend, ranging from 83.1% for logistic regression, 86.2% for random forest, and 89.3% for XGBoost on test datasets (**Figure 2**). Logistic regression achieved a sensitivity 78.2% and specificity 85.8%, suggesting balanced performance. Random forest increased specificity substantially (91.3%) but at the cost of slightly lower sensitivity (77.2%). XGBoost maintained the highest balance, with sensitivity 79.6% and specificity 94.8%, indicating it was most effective at identifying both unused and used appointments.

### Feature Odds Ratio

As shown in **Table 2**, logistic regression identified several demographics, clinical, and scheduling factors associated with appointment non-attendance. Asian patients had markedly higher odds of non-attendance compared to White patients, followed by Hispanic/Latino, Black, and those with other or unknown race. Female sex was associated with slightly lower odds compared to males. Medicaid and “Other” insurance were strongly protective compared with commercial insurance; self-pay and Medicare patients also had slightly reduced odds.

**Table 2.**
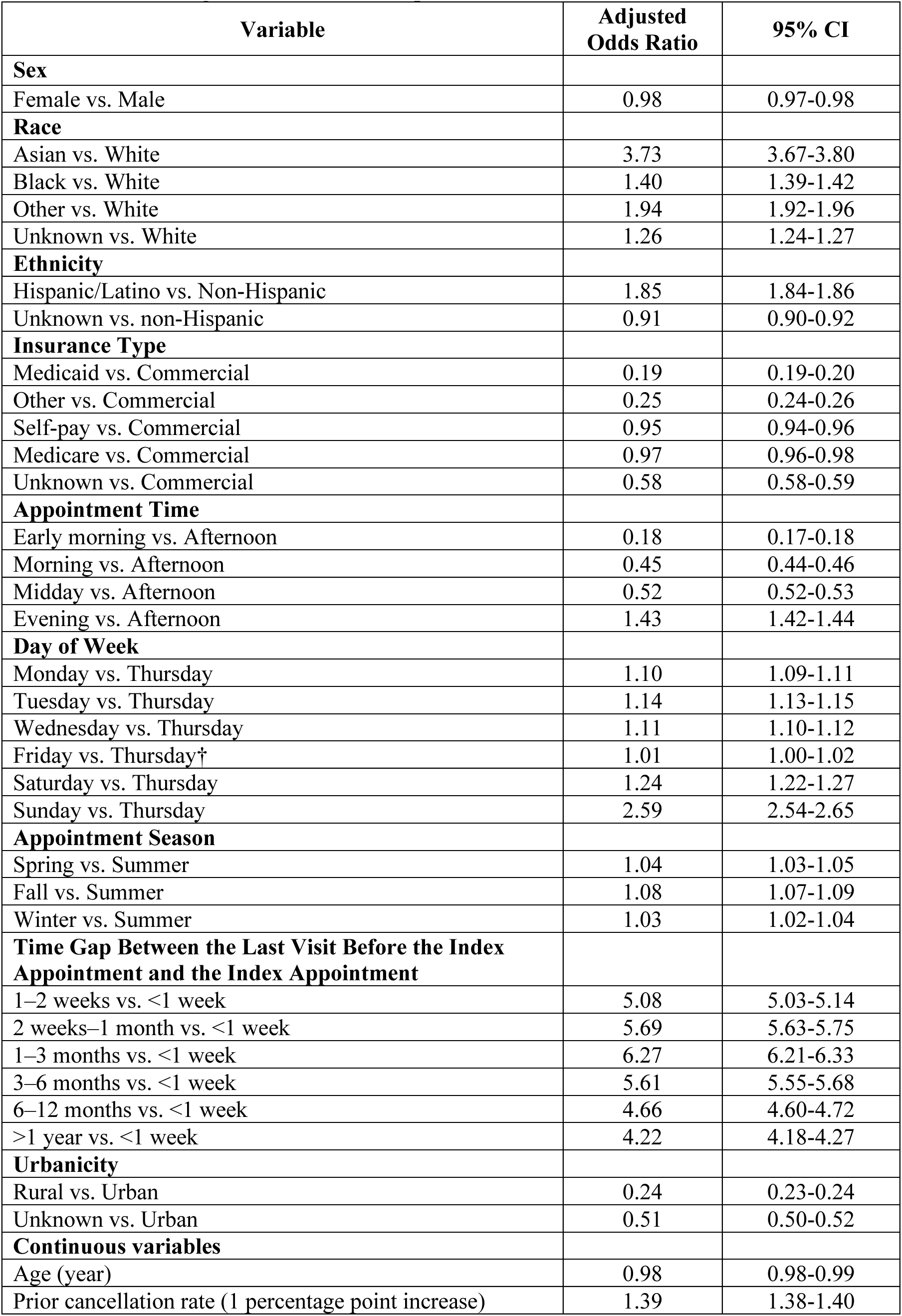
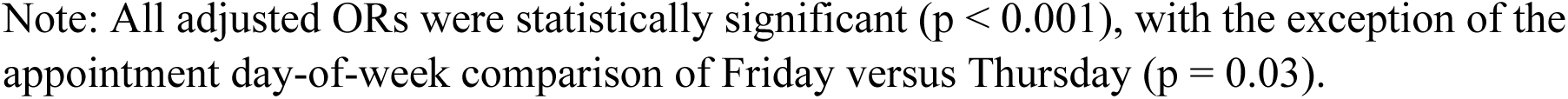
Results from regression model development.

| Variable | Adjusted Odds Ratio | 95% CI |
| --- | --- | --- |
| **Sex** |  |  |
| Female vs. Male | 0.98 | 0.97-0.98 |
| **Race** |  |  |
| Asian vs. White | 3.73 | 3.67-3.80 |
| Black vs. White | 1.40 | 1.39-1.42 |
| Other vs. White | 1.94 | 1.92-1.96 |
| Unknown vs. White | 1.26 | 1.24-1.27 |
| **Ethnicity** |  |  |
| Hispanic/Latino vs. Non-Hispanic | 1.85 | 1.84-1.86 |
| Unknown vs. non-Hispanic | 0.91 | 0.90-0.92 |
| **Insurance Type** |  |  |
| Medicaid vs. Commercial | 0.19 | 0.19-0.20 |
| Other vs. Commercial | 0.25 | 0.24-0.26 |
| Self-pay vs. Commercial | 0.95 | 0.94-0.96 |
| Medicare vs. Commercial | 0.97 | 0.96-0.98 |
| Unknown vs. Commercial | 0.58 | 0.58-0.59 |
| **Appointment Time** |  |  |
| Early morning vs. Afternoon | 0.18 | 0.17-0.18 |
| Morning vs. Afternoon | 0.45 | 0.44-0.46 |
| Midday vs. Afternoon | 0.52 | 0.52-0.53 |
| Evening vs. Afternoon | 1.43 | 1.42-1.44 |
| **Day of Week** |  |  |
| Monday vs. Thursday | 1.10 | 1.09-1.11 |
| Tuesday vs. Thursday | 1.14 | 1.13-1.15 |
| Wednesday vs. Thursday | 1.11 | 1.10-1.12 |
| Friday vs. Thursday† | 1.01 | 1.00-1.02 |
| Saturday vs. Thursday | 1.24 | 1.22-1.27 |
| Sunday vs. Thursday | 2.59 | 2.54-2.65 |
| **Appointment Season** |  |  |
| Spring vs. Summer | 1.04 | 1.03-1.05 |
| Fall vs. Summer | 1.08 | 1.07-1.09 |
| Winter vs. Summer | 1.03 | 1.02-1.04 |
| **Time Gap Between the Last Visit Before the Index Appointment and the Index Appointment** |  |  |
| 1–2 weeks vs. <1 week | 5.08 | 5.03-5.14 |
| 2 weeks–1 month vs. <1 week | 5.69 | 5.63-5.75 |
| 1–3 months vs. <1 week | 6.27 | 6.21-6.33 |
| 3–6 months vs. <1 week | 5.61 | 5.55-5.68 |
| 6–12 months vs. <1 week | 4.66 | 4.60-4.72 |
| >1 year vs. <1 week | 4.22 | 4.18-4.27 |
| **Urbanicity** |  |  |
| Rural vs. Urban | 0.24 | 0.23-0.24 |
| Unknown vs. Urban | 0.51 | 0.50-0.52 |
| **Continuous variables** |  |  |
| Age (year) | 0.98 | 0.98-0.99 |
| Prior cancellation rate (1 percentage point increase) | 1.39 | 1.38-1.40 |

Note: All adjusted ORs were statistically significant ( $p < 0.001$ ), with the exception of the appointment day-of-week comparison of Friday versus Thursday ( $p = 0.03$ ).

Appointments in early morning, morning, and midday had lower non-attendance risk than afternoon visits, while evening appointments increased risk. Sunday and Saturday appointments had significantly higher cancellation odds than Thursday. Fall appointments had slightly higher risk than summer.

Longer time gap since last visit strongly predicted unused appointments, peaking at 1–3 months. Rural patients were less likely to miss appointments than urban ones. Prior rate of unused appointments was also a strong predictor of future non-attendance.

These findings highlight that both visit characteristics (e.g., appointment day, appointment time, time gap), patient and clinical/behavioral characteristics (e.g., race/ethnicity, insurance type, prior behavior) contribute to the risk of missed appointments.

### Feature impact score

**Figure 3** shows the mean and SD of the impact scores of each feature based on the best-performing ML models, XGBoost model.

**Figure 3:**
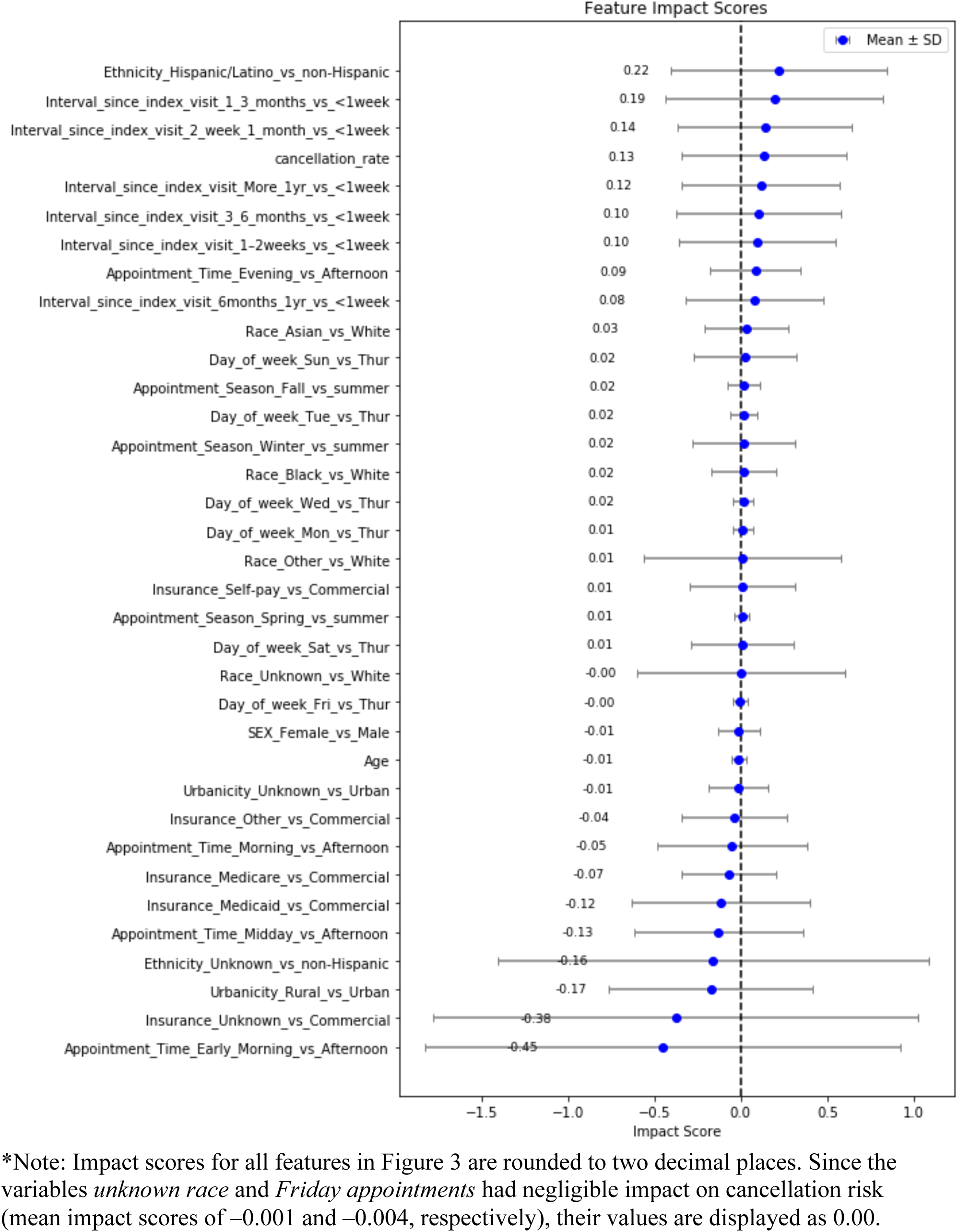
Feature Impact Score Analysis.

The very large standard deviation (SD) of the top five predictors suggests a heterogeneous (non-linear) association with the risk of unused appointments. These predictors are early morning vs. afternoon appointment, unknown insurance vs. commercial, unknown ethnicity vs. non-Hispanic, Hispanic or Latino ethnicity vs. non-Hispanic, and a time gap of 1–3 months vs. less than 1 week. The top five predictors showing a more homogeneous (linear) association are age, Friday vs. Thursday, spring vs. summer, Monday vs. Thursday, and Wednesday vs. Thursday.

## Discussion

In this large, nationally representative analysis, we found that ML models, particularly XGBoost, predicted unused outpatient appointments accurately, achieving an AUC of 0.95 with high accuracy, sensitivity, and specificity. These results suggest that ML, a key AI technology, can capture the complex interplay of patient and scheduling-related features of unused appointments. Notably, while prior studies using machine learning approaches have generally reported moderate to excellent AUCs in the range of 0.70 to 0.89^16–18^, our model reached an even higher AUC of 0.95. Although differences in study populations, data sources, and modeling strategies limit direct comparisons, the breadth and scale of our dataset may have contributed to a more robust predictive performance.

The feature impact from XGBoost, assessed using the population-level mean impact score, showed strong alignment with the odds ratios derived from logistic regression models. The variability in individual-level impact scores, captured by the SD, reflected the heterogeneity in how predictors were associated with unused appointments across individuals. Notably, even for predictors with a high mean impact score, some individuals exhibited a reduced risk of non-completion while others showed an increased risk for the same feature. This underscores the importance of personalized risk prediction. These findings highlight the advantage of ML in capturing heterogeneous, individual-level risk patterns. This heterogeneity suggests that appointment behavior is not uniform, even within groups traditionally considered high-risk.

Interpretability remains a critical factor for the adoption of AI-based tools in healthcare. Our use of explainable AI methods allowed us to quantify feature impact at both the population and individual level, providing transparency about why certain predictions were made. Such approaches, including the SHAP-inspired feature impact score, enhance trust by making complex models more clinically interpretable. Nonetheless, additional work is needed to determine whether these explanations are sufficient for clinicians to act on, and whether they ultimately improve provider decision-making and patient outcomes^25,26^.

Our findings align with prior studies that have identified risk factors for missed appointments, including minority race and ethnicity, younger age, longer scheduling lead times, and a history of prior cancellations^6,8,13^. However, Medicaid insurance status was associated with a reduced risk of missed appointments, which is inconsistent with findings reported in previous studies^6,13^. The reason behind this is not entirely clear, though our dataset is clearly more comprehensive. The inverse association with Medicaid coverage, while inconsistent with prior literature, may reflect differences in patient behavior or system-level policies that influence rescheduling. In our study, the outcome of missing appointments included both rescheduling and no-shows. This may reflect that patients with commercial insurance generally have more provider options and greater scheduling flexibility, making it easier for them to reschedule appointments^27^.

At the same time, this work extends the literature in several ways. Unlike studies confined to single institutions, specific specialities, or regional datasets^11,15–17,28,29^, our analysis drew on one of the largest U.S. national datasets available, enhancing generalizability across diverse populations and settings. A second contribution lies in the comparative approach: we evaluated both statistical and ML methods, showing that logistic regression provided interpretable insights into predictors while AI delivered superior predictive accuracy and captured non-linear relations. Finally, by reporting comprehensive performance measures, including calibration and discrimination, we provide a more complete evaluation than some prior works that focused solely on identifying risk factors.

Predictive models could inform more efficient and equitable scheduling strategies. Patients at higher predicted risk might benefit from earlier or midday appointment times, additional reminders, or telehealth alternatives, which have consistently shown lower absenteeism compared with in-person visits^30,31^. At the system level, predictions could be used to inform flexible scheduling strategies, such as selective overbooking or adjustment of staffing patterns, with the aim of reducing wasted time and improving patient flow. Equally important, the model underscores patterns of missed care that disproportionately affect certain demographic groups, such as Hispanic patients. Identifying these variances may help guide targeted interventions that extend beyond efficiency to strengthen access for populations that face persistent barriers to care.

While predictive models can improve efficiency, their use must also be carefully evaluated for potential discriminatory concerns. If patients at higher predicted risk of unused appointments are deprioritized or selectively overbooked, such approaches could inadvertently reinforce disparities for groups already experiencing structural barriers to care. To avoid this, predictive analytics should be paired with other interventions such as transportation support, flexible scheduling, and expanded telehealth access. Implementing these safeguards can help ensure that predictive modeling not only optimizes clinic operations but also strengthens access and fairness in care delivery^32,33^.

Finally, it is important to note that due to the large sample size used in this analysis, small differences between groups may reach statistical significance without necessarily reflecting clinically meaningful differences. While our models achieved strong predictive performance, including statistically significant associations, these findings should be interpreted in the context of their practical or clinical relevance. Randomized controlled trials should explore the real-world impact of interventions, based on these predictions, on patient outcomes and care delivery.

Overall, these findings should be interpreted with caution given the observational design, potential residual confounding, and the use of multiple modeling approaches. While the models demonstrated strong predictive performance, the results reflect associations rather than causal relationships and should be considered in the context of existing literature and study limitations. Given the use of a large, national EHR dataset, these findings are likely generalizable to similar healthcare systems in the United States, although applicability to settings with different healthcare structures or patient populations may be limited.

### Strengths and Limitations

This study has several strengths. It leveraged a large, nationally representative EHR dataset, enhancing the generalizability of findings across diverse populations and healthcare settings. Additionally, the use of both traditional statistical methods and advanced machine learning approaches allowed for robust modeling and comparison of predictive performance, while incorporating a wide range of patient, clinical, and scheduling factors.

This study has several limitations. First, the inability to distinguish between appointments that were canceled, missed entirely, or rescheduled may have introduced outcome misclassification, potentially biasing effect estimates toward the null and underestimating true associations.

Second, although the dataset was large and nationally representative, it did not include several potentially important variables, such as language preference, employment status, transportation access, or scheduling lead time. The absence of these factors may have resulted in residual confounding, potentially affecting both the magnitude and direction of observed associations.

Third, reliance on EHR data introduces the possibility of measurement error and misclassification of predictors, which may attenuate associations and slightly reduce model performance, as predictive models are highly dependent on data quality.^34^ Data quality issues, including potential inaccuracies in the underlying database, may also have influenced results. Fourth, we analyzed one random encounter per patient to preserve independence, which limited the ability to assess longitudinal patterns such as repeated cancellations. This approach may underestimate the impact of prior behavior over time. Fifth, while machine learning models such as XGBoost demonstrated strong predictive performance, they may be less transparent than traditional statistical approaches, which could affect clinician trust and implementation in practice.

Finally, the retrospective design limits causal inference, and prospective evaluation in real-world scheduling systems is needed to determine whether these predictive models improve operational efficiency and patient outcomes. In addition, due to limited documentation of no-shows (or left-before-seen encounters, <0.1% of visits), sensitivity analyses specifically targeting no-show risk were not feasible. Although the large sample size increases statistical power, it may also result in statistically significant findings that are not clinically meaningful. Nonetheless, given the consistency of findings across models and the scale of the dataset, these limitations are unlikely to substantially alter the overall conclusions.

## Conclusions

In summary, this study demonstrates that missed and cancelled outpatient appointments can be accurately predicted using routinely collected data from a large, nationally representative EHR repository. By leveraging advanced machine learning methods, particularly XGBoost, we achieved strong predictive performance and revealed non-linear, individualized associations between patient and appointment characteristics and the likelihood of appointment non-completion.

Beyond predictive accuracy, our findings emphasize the importance of explainable and personalized modeling. The observed heterogeneity in predictor impact underscores that appointment behavior is not uniform, even within traditionally high-risk groups. Integrating such interpretable models into clinical scheduling workflows could enable more proactive and equitable interventions. For example, offering alternative times, telehealth options, or transportation support for those at highest risk of missed visits.

Future work should focus on prospective validation within live scheduling systems to assess whether predictive models meaningfully improve operational efficiency, patient outcomes, and access to care. Equally important, their implementation must be guided by ethical and equity considerations to ensure that data-driven approaches reduce, rather than reinforce, disparities in healthcare delivery.

## Supporting information

Supplementary File 1: STROBE checklist

## Data availability statement

The data that support the findings of this study are available from the Cerner Real World Database (Oracle Health). These data are not publicly available due to licensing restrictions but may be accessed by qualified researchers with permission from Oracle Health. The statistical code used in this study is available from the corresponding author upon reasonable request.

## Author Statement

**Seyedeh Azadeh Miran:** Conceptualization, study design, data acquisition, formal analysis, machine learning modeling, interpretation of results and drafting of the manuscript.

**Yan Cheng:** Statistical analysis support, interpretation of results, and drafting and critical revision of the manuscript.

**Charles Faselis:** Critical revision of the results and the manuscript

**Cynthia Brandt:** Critical revision of the results and the manuscript

**Sean Vasaitis:** Study design, results review, critical revision of the manuscript

**LaQuandra Nesbitt:** Study design, results review

**Linda Zanin:** Conceptualization, participation in discussion

**Senait Tekle:** Participation in discussion, result review

**Ali Ahmed:** Critical review of methodology and results

**Stuart J. Nelson:** Conceptualization, study design, methodology and critical revision of the manuscript

**Qing Zeng-Treitler:** Conceptualization, study design, methodology and critical revision of the manuscript.

All authors contributed to drafting or revising the manuscript, approved the final version for publication, and agree to be accountable for all aspects of the work.

## Acknowledgements

None.

## Authorship

All authors had access to data output and had roles in the writing of the manuscript.

