## Supplementary File 1: STROBE checklist for "Nationwide Prediction of Missed and Cancelled Appointments Using Real-World EHR Data"

| | Item Description | Location (or reason for not reporting) |
|---|---|---|
| **Title and abstract** | | |
| 1a. Indicate the study's design | Indicate the study's design with a commonly used term in the title or the abstract. | Title + Abstract (Design: Retrospective observational study) |
| 1b. Abstract | Provide in the abstract an informative and balanced summary of what was done and what was found. | Structured Abstract (Objectives, Design, Setting, Participants, Methods, Results, Conclusions) |
| **Introduction** | | |
| 2. Background / rationale | Explain the scientific background and rationale for the investigation being reported. | Introduction, paragraphs 1–3 |
| 3. Objectives | State specific objectives, including any prespecified hypotheses. | Introduction, final paragraph + Abstract (Objectives section) |
| **Methods** | | |
| 4. Study design | Present key elements of study design early in the paper. | Methods – Study Design + Abstract (Design) |
| 5. Setting | Describe the setting, locations, and relevant dates, including periods of | Methods – Data Source + Abstract (Setting: Cerner |

| | | |
|---|---|---|
| | recruitment, exposure, follow-up, and data collection. | database, 2010–2025, U.S.) |
| 6a. Eligibility criteria | **Cohort study:** Give the eligibility criteria, and the sources and methods of selection of participants. Describe methods of follow-up. **Case-control study:** Give the eligibility criteria, and the sources and methods of case ascertainment and control selection. Give the rationale for the choice of cases and controls. **Cross-sectional study:** Give the eligibility criteria, and the sources and methods of selection of participants. | Methods – Study Design + Abstract (Participants) |
| 6b. Matching criteria | **Cohort study:** For matched studies, give matching criteria and number of exposed and unexposed. **Case-control study:** For matched studies, give matching criteria and the number of controls per case. | Not applicable |
| 7. Variables | Clearly define all outcomes, exposures, predictors, potential confounders, and effect modifiers. Give diagnostic criteria, if applicable. | Methods – Outcome Variable + Predictor Variables + Abstract (Outcome + Methods) |
| 8. Data sources / measurement | For each variable of interest give sources of data and details of methods of assessment (measurement). Describe comparability of assessment methods if there is more than one group. | Methods – Data Source + Predictor Variables |
| 9. Bias | Describe any efforts to address potential sources of bias. | Methods – Study Design (random selection to reduce selection bias and data leakage);<br><br>Discussion – Limitations (residual confounding, misclassification, and |

| | | |
|---|---|---|
| | | measurement error) |
| 10. Study size | Explain how the study size was arrived at. | Methods – Study Design (selection of one encounter per patient);<br>Results – Cohort and Encounter Selection (final sample size n = 5,699,861) |
| 11. Quantitative variables | Explain how quantitative variables were handled in the analyses. If applicable, describe which groupings were chosen, and why. | Methods – Predictor Variables (categorization described) |
| 12a. Statistical methods | Describe all statistical methods, including those used to control for confounding. | Methods – Statistical Analysis + Abstract (Methods) |
| 12b. Statistical methods – subgroups and interactions | Describe any methods used to examine subgroups and interactions. | Not performed |
| 12c. Statistical methods – missing data | Explain how missing data were addressed. | Methods – Statistical Analysis (missing data handling: insurance type categorized as "unknown") |
| 12di. Statistical methods – loss to follow-up | **Cohort study:** If applicable, describe how loss to follow-up was addressed. | Not applicable |
| 12dii. Statistical methods – matching cases and controls | **Case-control study:** If applicable, explain how matching of cases and controls was addressed. | Not applicable |

| | | |
|---|---|---|
| 12diii. Statistical methods – sampling strategy | **Cross-sectional study:** If applicable, describe analytical methods taking account of sampling strategy. | Methods – Study Design (random selection of one outpatient encounter per patient to ensure independence and reduce bias) |
| 12e. Statistical methods – sensitivity analyses | Describe any sensitivity analyses. | Not performed; sensitivity analysis for missed appointments was not feasible due to limited documentation in the database, as described in the Limitations section. |
| **Results** | | |
| 13a. Participant numbers | Report the numbers of individuals at each stage of the study—e.g., numbers potentially eligible, examined for eligibility, confirmed eligible, included in the study, completing follow-up, and analysed; Consider use of a flow diagram. | Results – Cohort and Encounter Selection , Abstract (Participants + Results), Figure 1 |
| 13b. Participants – non-participation | Give reasons for non-participation at each stage. | Results – Cohort and Encounter Selection, Figure 1 |
| 13c. Participants – flow diagram | Consider the use of a flow diagram. | Figure 1 |
| 14a. Descriptive data – participant characteristic s | Give characteristics of study participants (e.g., demographic, clinical, social) and information on exposures and potential confounders. Present the information in a table. | Table 1 |
| 14b. Descriptive data – missing data | Indicate the number of participants with missing data for each variable of interest. | Table 1 (insurance type "unknown" category); Methods – Missing data handling |
| 14c. Descriptive data – follow-up time | **Cohort study:** Summarise follow-up time—e.g., average and total amount. | Not applicable |

| | | |
|---|---|---|
| 15. Outcome data | **Cohort study:** Report numbers of outcome events or summary measures over time. **Case-control study:** Report numbers in each exposure category, or summary measures of exposure. **Cross-sectional study:** Report numbers of outcome events or summary measures. | Results – Characteristics and Appointment Cancellation |
| 16a. Main results | Give unadjusted estimates and, if applicable, confounder-adjusted estimates and their precision (e.g., 95% confidence intervals). Make clear which confounders were adjusted for and why they were included. | Results – Multivariable logistic regression analysis (adjusted odds ratios with 95% confidence intervals), Table 2; Results – Feature Odds Ratio section |
| 16b. Main results – category boundaries | Report category boundaries when continuous variables were categorised. | Methods – Predictor Variables, Table 1 |
| 16c. Main results – risk | If relevant, consider translating estimates of relative risk into absolute risk for a meaningful time period. | Not applicable |
| 17. Other analyses | Report other analyses done—e.g., analyses of subgroups and interactions, and sensitivity analyses. | No additional subgroup or sensitivity analyses were performed; this is discussed in the Limitations section. |
| **Discussion** | | |
| 18. Key results | Summarise key results with reference to study objectives. | Discussion, paragraph 1 |
| 19. Limitations | Discuss limitations of the study, taking into account sources of potential bias or imprecision. Discuss both direction and magnitude of any potential bias. | Discussion – Limitations section |
| 20. Interpretation | Give a cautious overall interpretation considering objectives, limitations, multiplicity of analyses, results from similar studies, and other relevant evidence. | Discussion – Interpretation paragraphs (including final paragraph) |
| 21. Generalisability | Discuss the generalisability (external validity) of the study results. | Discussion – Generalisability (national dataset, |

| | | |
|---|---|---|
| | | comparison with prior studies, and applicability across diverse populations and settings) |
| **Other information** | | |
| 22. Funding | Give the source of funding and the role of the funders for the present study and, if applicable, for the original study on which the present article is based. | Title page (NIH AIM-AHEAD grant and VA resources; role of funders stated) |
